# MPSE provides fast, flexible, and efficient means to identify newborns who will benefit from whole genome sequencing within the first 48 hours of NICU admission

**DOI:** 10.1101/2024.11.05.24316150

**Authors:** Bennet Peterson, Edwin F. Juarez, Barry Moore, Edgar Javier Hernandez, Erwin Frise, Jianrong Li, Yves Lussier, Martin Tristani-Firouzi, Martin G. Reese, Sabrina Malone Jenkins, Stephen F. Kingsmore, Matthew N. Bainbridge, Mark Yandell

**Affiliations:** Department of Biomedical Informatics, University of Utah, Salt Lake City, UT, USA; Rady Children’s Institute for Genomic Medicine, San Diego, CA, USA; Department of Human Genetics, Utah Center for Genetic Discovery, University of Utah, Salt Lake City, UT, USA; Fabric Genomics Inc., Oakland, CA, USA; Division of Pediatric Cardiology, University of Utah School of Medicine, Salt Lake City, UT, USA; Division of Neonatology, Department of Pediatrics, University of Utah School of Medicine, Salt Lake City, UT, USA

## Abstract

**Background:** Identifying patients who would benefit from whole genome sequencing (WGS) is difficult and time-consuming due to complex eligibility criteria, lack of neonatologist familiarity with WGS ordering, and evolving clinical features. In previous work, we showed that MPSE, the Mendelian Phenotype Search Engine, can provide automated prioritization of probands for WGS while maintaining current diagnostic rates. MPSE is now in use in multiple hospital networks, but questions still surround how to best prioritize patients for WGS.

**Methods:** Here we use the clinical histories of 2,885 neonatal intensive care unit (NICU) admits from two institutions to explore further questions regarding how to best prioritize NICU admits for WGS. First, we ask if changes to the machine learning (ML) classifier and the clinical natural language processing (CNLP) tools used for generating patient phenotype descriptions might improve MPSE’s performance. Second, we explore the utility of using alternative data types as inputs to MPSE. Lastly, we conduct a longitudinal analysis of MPSE’s ability to identify probands for WGS.

**Results:** Eight different ML classifiers, five CNLP tools, and four previously untested alternative data types were used to train and validate MPSE models. MPSE achieved high predictive performance across multiple classifiers (max AUC=0.93), CNLP tools (max AUC=0.91), and input data types (max AUC=0.91). Longitudinal analysis of MPSE scores revealed a significant separation between cases/controls and diagnostic/non-diagnostic cases within 48 hours of NICU admission.

**Conclusions:** MPSE provides a highly flexible and portable framework for automated prioritization of critically ill newborns for WGS. We find that MPSE’s performance is largely agnostic with respect to CNLP tools. Moreover, structured data such as ICD codes can serve as an effective alternative input to MPSE when access to clinical notes or CNLP pipelines is problematic. Finally, MPSE can identify children most likely to benefit from WGS within 48 hours of admission to the NICU, a critical window for maximally impactful care.

## Background

Each year approximately 7 million infants worldwide are born with genetic disorders. Many are diagnosed and treated in the neonatal intensive care unit (NICU)^1^. Rapid progression of disease in acutely ill infants necessitates equally rapid diagnosis to implement personalized interventions. In recent years, whole genome sequencing (WGS) has emerged as a primary diagnostic tool^2–4^. An estimated one-fifth of NICU admissions involve Mendelian diseases, with WGS diagnostic yield commonly in the range of 25-50%^5–8^. However, identifying infants for WGS is difficult and time-consuming due to complex eligibility criteria, lack of neonatologist familiarity with WGS ordering, and evolving clinical features.

Manual review and prioritized selection of patient phenotypes is a time-consuming and expensive process, hindering WGS application in the NICU^9,10^. Since interpretation is phenotype-driven, incomplete or erroneous phenotype selection can result in false negative results. Failure to adhere to payer eligibility criteria can lead to refusal of reimbursement. Complicating this are the complexity of eligibility criteria and differences between payers. Recent efforts explore clinical natural language processing (CNLP) to automatically generate Human Phenotype Ontology (HPO)-based phenotype descriptions from clinical notes, and have demonstrated diagnostic rates comparable to manual methods^9,11^. Automation promises scalability and efficiency in patient triage for sequencing. In previously published work, we have shown that combining CNLP with a machine learning-based prioritization tool, the Mendelian Phenotype Search Engine (MPSE) provides effective means to prioritize patients for WGS using electronic health records (EHRs)^12,13^.

Perhaps the greatest benefit from tools like MPSE will be seen by limited resource healthcare systems which may lack the expertise, funding, or data necessary to develop in-house computational frameworks for driving genomics-based clinical care. Generalizability and adaptability are therefore essential. With these facts in mind, we have explored MPSE’s utility across multiple patient populations, data sources, and input data types. Our results reveal that MPSE is fast, flexible, generalizable, and highly portable.

Flexibility, generalizability, and portability are important characteristics of MPSE, but time to WGS order is of critical importance in the NICU clinical setting. Earlier identification of patients likely to benefit from WGS, ideally as soon as possible after NICU admission, can significantly enhance care by enabling earlier disease diagnosis and timelier, more personalized interventions^2,14,15^. Here we show that MPSE can identify those children most likely to benefit from WGS within the first 24 hours of admission to the NICU, a critical window for maximally impactful care. Moreover, we find that the MPSE scores of patients who are ultimately diagnosed with Mendelian diseases are higher than those of sequenced but non-diagnostic cases, a statistically significant trend that appears at 48 hours post-admission and continues across the entire duration of the NICU stay. These findings argue for MPSE’s use as a proactive monitoring tool throughout the NICU stay.

## Methods

### Datasets

Our clinical cohort comprised 293 probands who underwent rapid whole genome sequencing (rWGS) at Rady Children’s Hospital in San Diego (RCHSD), 85 of whom received a molecular diagnosis for a Mendelian disorder. These 293 individuals were selected from symptomatic children enrolled in prior studies exploring the diagnostic rate, time to diagnosis, clinical utility, outcomes, and healthcare utilization of rWGS between July 26, 2016, and September 25, 2018, at RCHSD (ClinicalTrials.gov identifiers: NCT03211039, NCT02917460, and NCT03385876). All participants presented with symptomatic illnesses of unknown etiology and suspected genetic disorders. The diagnosed cases provide a real-world population displaying various Mendelian conditions arising from diverse modes of disease inheritance and disease-causing genotypes. An additional 756 patients admitted to the NICU at RCHSD in 2018 were also included. These patients were added to enrich the dataset with a broader spectrum of phenotypes not necessarily associated with Mendelian diseases. In total, the RCHSD dataset used in this study contains a total of 1,049 individuals. Additional details are provided in^2,7,9,14,15^.

We also employed a second, independent dataset consisting of 1,838 newborn patients admitted to the University of Utah level III NICU from January 2020 to December 2022, the approximate study period of the Utah NeoSeq Project. The Utah NeoSeq Project was a multidisciplinary, longitudinal rapid genome sequencing program conducted at the University of Utah to improve genetic diagnosis in critically ill infants in the NICU. Within the Utah cohort of 1,838 patients, 65 were selected for rWGS based on manual chart review as part of the NeoSeq study. 26 of these children received a molecular diagnosis.

### Statistical classifiers

Various Machine Learning (ML) classifiers were used to train multiple independent MPSE models. The original MPSE algorithm employed a Naive Bayes classifier and is described in detail in our proof-of-concept work^12^. Additional MPSE models were trained using the following ML classifiers: K-Nearest Neighbors (KNN), Decision Trees (DT), Random Forests (RF), Logistic Regression (LR), Gradient Boost Machine (GBM), Support Vector Machine (SVM), and Multi-Layer Perceptron (MLP). Each classifier was implemented using *scikit-learn*, a general-purpose machine learning library written in the Python programming language^16^. Each method was run with scikit-learn version 1.4.2 default parameters. Models were trained using the RCHSD cohort (n=1,049) and internally validated using stratified K-fold cross validation (K=8). Each of the trained classifiers was subsequently validated externally with five randomly split subsets of the Utah cohort (n=419 per set).

### Phenotype and alternative data types

Highly curated, manually created HPO-based phenotype descriptions were provided for each of the 65 sequenced University of Utah probands. CNLP-derived phenotype descriptions were generated for all 1,049 RCHSD probands and the 1,838 University of Utah probands by CNLP analysis of clinical notes recorded during NICU stay using CLiX^17^. Additional CNLP-derived phenotype descriptions were generated using the following text mining and CNLP methods: ClinPhen^18^, cTAKES^19^, MedLEE^20^, and MetaMapLite^21^ for the University of Utah probands. CLiX and ClinPhen generate HPO terms directly from clinic notes, while cTAKES, MedLEE, and MetaMapLite return Unified Medical Language System (UMLS)^22^ Concept Unique Identifiers (CUIs) which were then mapped to HPO terms using the UMLS Metathesaurus’ network hierarchy. For sequenced patients, clinical notes dated post-WGS were excluded from analysis to remove notes containing sequencing results. In addition to CNLP-derived phenotype data, ICD-10 diagnosis codes, laboratory tests, medications, and other hospital orders were collected for all University of Utah probands to serve as alternative data sources for MPSE modeling. Unlike free-text clinic notes, these alternative data types are stored in structured form within the University of Utah’s Enterprise Data Warehouse (EDW) and were extracted using automated database queries. Laboratory tests and medication orders were encoded as binary variables to represent the presence/absence of a test or order without the context of the test result or medication dosage, frequency, adherence, etc..

### Calculating semantic similarity between phenotype sets

We calculated pairwise semantic similarity between physician-and CNLP-generated HPO term sets for patients using the Python package PyHPO v3.1.4. PyHPO calculates the similarity between two HPO sets as described in^23,24^. We also generated simulated, ‘randomized’, HPO term sets for every proband in order to provide a “null” distribution for our semantic similarity calculations. For each “real” pairwise set comparison, simulated HPO sets of equal size were randomly sampled from the HPO. Semantic similarity between these sets of randomly sampled terms was then calculated using the same method that was used to compare the manual and CNLP sets to one another. Significant differences between “real” and “randomized” set similarities were tested for using paired Student’s T-test.

### Testing the precision and diagnostic yield of MPSE

We calculated the precision and diagnostic yield at K for the 5, 10, 25, 45, and 65 top-scoring patients by dividing the number of true positives by K. For calculating precision, a true positive was defined as a WGS-selected patient that was classified (flagged) by MPSE as a WGS candidate based upon the contents of its clinical notes using a particular C-NLP tool, i.e., its MPSE score > 0. For calculating diagnostic yield, a true positive was defined as a WGS-diagnosed patient that was flagged by MPSE (MPSE score > 0).

### Gene prioritization using NLP-derived phenotype descriptions

Artificial Intelligence (AI)-based prioritization and scoring of candidate disease genes for the diagnosed probands was performed using Fabric GEM^11^. GEM is a commercial tool for AI-assisted clinical interpretation of WES and WGS. It has been licensed by the University of Utah and Rady Children’s Hospitals from Fabric Genomics Inc. Additional licensing information is available from Fabric Genomics Inc. GEM inputs are genetic variant calls in VCF format and case metadata, including parental affection status, and patient phenotypes in the form of HPO terms. For these analyses, GEM was run six times for each NeoSeq proband, varying only the input HPO lists, the first with the proband’s physician-selected terms and the remaining runs using HPO term sets created with the five different CNLP tools.

### Longitudinal analysis of MPSE scores

We conducted a longitudinal analysis of MPSE scores across each University of Utah patient’s NICU stay. For this analysis, we employed the original MPSE model trained on the RCHSD cohort and utilized CLiX-derived HPO phenotype descriptions. Human Phenotype Ontology (HPO) terms were timestamped according to the date of the clinical note, with each day’s cumulative HPO list being used for MPSE score calculations. Six sequenced University of Utah patients had to be excluded from this analysis because a data upload error left clinic note timestamps unavailable for these patients, leaving 59 remaining sequenced patients available for longitudinal analysis. Subsequently, we calculated average daily MPSE scores for unsequenced controls, sequenced but not diagnostic cases, and diagnostic cases. We estimated the daily probability MPSE would recommend patients from these groups for sequencing using a score threshold (calculated individually for each day) of 2 standard deviations above the mean score of unsequenced control patients, and calculated the associated hazards ratio with Cox proportional hazards regression analysis using the R *survival* package (v3.7.0).

### Comparing diagnostic yield between MPSE and ACMG practice guideline

We calculated the 10-day post-admission diagnostic yields achieved by MPSE versus that achieved using American College of Medical Genetics and Genomics (ACMG) criteria^25^. University of Utah cohort patients (n=1838) were selected for WGS by MPSE at day 10 using the score thresholding approach detailed in the longitudinal analysis section (above). Patients from the same group and time point were again selected for WGS according to the 2021 ACMG Practice Guideline^25^ for the use of exome and genome sequencing for pediatric patients with congenital anomalies or intellectual disability. This guideline strongly recommends clinical genome sequencing as a first or second-tier test for patients with congenital anomalies prior to one year of age. Thus, the selection criteria for ACMG was one or more HPO terms indicating a congenital anomaly. Because phenotype terms representing congenital anomalies are scattered throughout the HPO hierarchy and not related under a single top-level parent term, we manually gathered a list of 12 common and clinically impactful congenital anomalies: Neural tube defect (HP:0045005), Spina bifida (HP:0002414), Status epilepticus (HP:0002133), Hypotonia (HP:0001252), Hyperammonemia (HP:0001987), Arthrogryposis multiplex congenita (HP:0002804), Congenital hypothyroidism (HP:0000851), Abnormal heart morphology (HP:0001627), Abnormal EKG (HP:0003115), Congenital lactic acidosis (HP:0004902), Cleft lip (HP:0410030), and Cleft palate (HP:0000175). The proportion of selected individuals who were previously diagnosed by WGS was then calculated using these two methods.

## Results and discussion

In previous work^12,13^, we showed that MPSE provides scalable, automated means for prioritizing probands for WGS while maintaining diagnostic rates. Here we use the clinical histories of 2,941 NICU admits drawn from two different institutions to explore three outstanding questions regarding automated prioritization of NICU admits for WGS.

First, we explore how changes to the ML classifier and the CNLP tools used for generating patient phenotype descriptions affect prioritization accuracy. Second, we explore the utility of using additional data types as inputs to MPSE, such as laboratory test orders and ICD diagnostic and procedure codes, both as an adjunct to CNLP-derived (HPO) phenotype descriptions and as alternative inputs. Lastly, we conduct a longitudinal analysis of MPSE’s discriminative abilities, exploring its power to successfully identify probands for WGS within the first 24 to 48 hours of NICU admission, a critical window for maximally impactful care.

Our findings speak directly to the ability of the MPSE approach to assist with scalable and cost-effective democratization of WGS nationwide. We show that our approach performs well using a variety of commercial and open-source ML algorithms and CNLP tools. We also observe strong performance using either clinical notes and/or structured data in the form of ICD codes and laboratory test orders. Most importantly of all, we show that MPSE can identify those children most likely to benefit from WGS within the first 24 to 48 hours of their admission to the NICU, in some cases long before human case review led to the same conclusion.

### Naive Bayes and Support Vector Machines are well-suited for robust phenotype-driven patient prioritization

MPSE uses Bernoulli Naive Bayes (BNB) as its default classifier^12,26^. To test the hypothesis that other ML approaches might outperform Naïve Bayes, we created versions of MPSE using 7 additional classifiers: K-Nearest Neighbors (KNN)^27^, Decision Trees (DT)^28^, Random Forests (RF)^29^, Logistic Regression (LR)^30^, Gradient Boost Machine (GBM)^31^, Support Vector Machine (SVM)^32^, Multi-Layer Perceptron (MLP)^33^. All models were built using the scikit-learn^34^ Python package using default model parameters.

We used a two-tiered approach to evaluate each model’s performance: internally resampling the RCHSD dataset, followed by a second round of cross-validation using the orthogonal University of Utah dataset to assess transportability.

#### Cross-Validation

The first validation tier consisted of stratified 8-fold internal cross-validation using the RCHSD dataset. Stratified K-fold cross-validation splits the training data into K folds while ensuring that each fold has approximately the same proportion of cases and controls as the original dataset. Each fold serves as a test set for a model trained using the remaining K-1 folds. Stratifying the splits helps prevent the model from being biased towards the majority class. Performance statistics for the eight splits were averaged within each model and are presented in **Table 1**. The results of this resampling-based cross-validation using the RCHSD dataset show that sophisticated classifiers, such as Support Vector Machines and Multi-Layer Perceptrons, outperform Naive Bayes. K-Nearest Neighbors and Decision Trees proved inferior to BNB by most measures.

**Table 1.** More sophisticated MPSE models achieve high classification accuracy and outperform simpler ones within the training cohort. Averaged internal cross-validation performance metrics for different statistical classifiers. Each model was trained on the same RCHSD cohort (756 unsequenced controls; 293 sequenced cases) using stratified K-fold cross-validation with K=8. Table rows are ordered by increasing F1-score. Classifier abbreviations: Bernoulli Naive Bayes (BNB), K-Nearest Neighbors (KNN), Decision Trees (DT), Random Forests (RF), Logistic Regression (LR), Gradient Boost Machine (GBM), Support Vector Machine (SVM), Multi-Layer Perceptron (MLP). Precision = TP/(TP+FP); Recall = TP/(TP+FN); F1 = 2*TP/(2*TP+FP+FN); Accuracy = (TP+TN)/(TP+TN+FP+FN); AUROC = area under the receiver operator characteristic curve

| Rank | Classifier | Precision | Recall | F1 | Accuracy | AUROC |
| --- | --- | --- | --- | --- | --- | --- |
| 1 | SVM | 0.83 | 0.72 | 0.77 | 0.88 | 0.93 |
| 2 | MLP | 0.82 | 0.73 | 0.77 | 0.88 | 0.92 |
| 3 | LR | 0.81 | 0.73 | 0.76 | 0.88 | 0.92 |
| 4 | GBM | 0.85 | 0.60 | 0.70 | 0.86 | 0.90 |
| 5 | RF | 0.89 | 0.53 | 0.66 | 0.85 | 0.92 |
| 6 | BNB | 0.68 | 0.54 | 0.60 | 0.80 | 0.85 |
| 7 | DT | 0.64 | 0.58 | 0.60 | 0.79 | 0.72 |
| 8 | KNN | 0.76 | 0.11 | 0.20 | 0.74 | 0.68 |

#### Transportability Analysis

The second validation tier, external validation, consisted of using the 8 MPSE models trained with RCHSD data to make predictions on five randomly split subsets of the Utah dataset. Each model was applied to the same five external validation sets and performance statistics for the five sets were averaged and are presented in **Table 2**. The external validation results contrast with the internal validation results, with MPSE’s original model, Bernoulli Naive Bayes (BNB), being the top-performing approach. The BNB model was the only model that did not see a significant drop in prediction performance moving from internal to external validation; in fact, BNB exhibited an increase in F1-score, accuracy, and AUROC with external validation. The difference in relative performance between internal and external validations is likely caused by overfitting and the curse of dimensionality. Naive Bayes is known to be less susceptible to both^35,36^. Interestingly, the other models largely maintained their rank order between internal and external validation, except GBM, which dropped from rank 4 to rank 7, and BNB which jumped from rank 6 to rank 1. To summarize, some classifiers performed well on the training data (LR, MLP, and SVM), while some models are more transportable (SVM, BNB). Transportability is essential if MPSE is to be deployed in different health systems.

**Table 2.** MPSE’s Naive Bayes (BNB) model is the most robust ML technique by external validation. Averaged external validation performance metrics for different statistical classifiers. Each model was validated on the same five randomly split subsets of the University of Utah cohort (354 unsequenced controls per split; 65 sequenced cases per split). Rows are ordered by increasing F1-score. Precision = TP/(TP+FP); Recall = TP/(TP+FN); F1 = 2*TP/(2*TP+FP+FN); Accuracy = (TP+TN)/(TP+TN+FP+FN); AUROC = area under the receiver operator characteristic curve

| Rank | Classifier | Precision | Recall | F1 | Accuracy | AUROC |
| --- | --- | --- | --- | --- | --- | --- |
| 1 | BNB | 0.73 | 0.52 | 0.61 | 0.89 | 0.89 |
| 2 | SVM | 0.45 | 0.49 | 0.47 | 0.83 | 0.83 |
| 3 | MLP | 0.39 | 0.49 | 0.43 | 0.80 | 0.73 |
| 4 | LR | 0.35 | 0.48 | 0.41 | 0.78 | 0.71 |
| 5 | RF | 0.57 | 0.29 | 0.38 | 0.86 | 0.79 |
| 6 | DT | 0.24 | 0.49 | 0.32 | 0.68 | 0.60 |
| 7 | GBM | 0.20 | 0.49 | 0.28 | 0.61 | 0.59 |
| 8 | KNN | 1.00 | 0.02 | 0.03 | 0.85 | 0.54 |

### Comparing clinical NLP tool outputs

All phenotype data used in our initial publications^12,13^ was generated from patient clinic notes using the CNLP software CLiX. CLiX is a proprietary clinical NLP technology developed by the commercial healthcare analytics company Clinithink^17^. Given the ultimate goal of developing MPSE for adoption by diverse hospital and clinic systems, we sought to determine MPSE’s performance using phenotype data produced by other tools as well. We conducted a series of analyses on 5 different CNLP tools to compare their relative utility for use with MPSE: ClinPhen^18^, CLiX, cTAKES^19^, MedLEE^20^, and MetaMapLite^21^. A brief description of these tools is given in **Additional file 1: Table S1**. Before assessing MPSE’s performance using phenotype data produced by these different CNLP tools, we first compared the phenotype descriptions (HPO term sets) generated by these tools using the same sets of clinic notes --in this case, the notes from 1,838 University of Utah NICU admits.

#### Term Counts

Summary statistics for unique HPO term counts generated by each CNLP tool as well as the “manual” term sets identified by expert physicians are given in **Additional file 1: Table S2**. The HPO term sets used throughout this work were pre-processed by removing parent terms to keep only the most specific phenotype terms. In every case, the CNLP tools all produced larger HPO term sets per patient than did expert review. Among HPO term sets for the University of Utah NeoSeq patients, MedLEE yielded the fewest terms (average 31.4 terms per patient) while CLiX yielded the most terms (average 111.2 terms per patient), nearly twice as many as the next most prolific tool cTAKES (70.5 terms per patient). Unsequenced University of Utah NICU patients had significantly fewer terms in their phenotype descriptions than NeoSeq patients, consistent with our observations from other patient cohorts^12^.

#### Semantic Similarity

We also calculated pairwise semantic similarity coefficients across all the NeoSeq phenotype sets. Semantic similarity is different from strict identity-based similarity measures, such as unweighted Jaccard similarity, in that two terms can be non-identical but still contribute positively to the similarity coefficient if they are neighbors, i.e., they lie near one another in the HPO directed acyclic graph. Note, however, that due to the nature of semantic similarity, any two arbitrary term sets sampled from the HPO will have a semantic similarity above zero even if no terms are shared between the sets. To estimate the probability that the semantic similarities of the term sets produced by the tools are statistically different from a null or random distribution, for each pairwise comparison, we sampled the HPO to yield two random term sets with sizes identical to the original sets. Semantic similarity coefficients were then calculated for these randomized sets and plotted alongside the real data in **Figure 1**.

**Figure 1.**
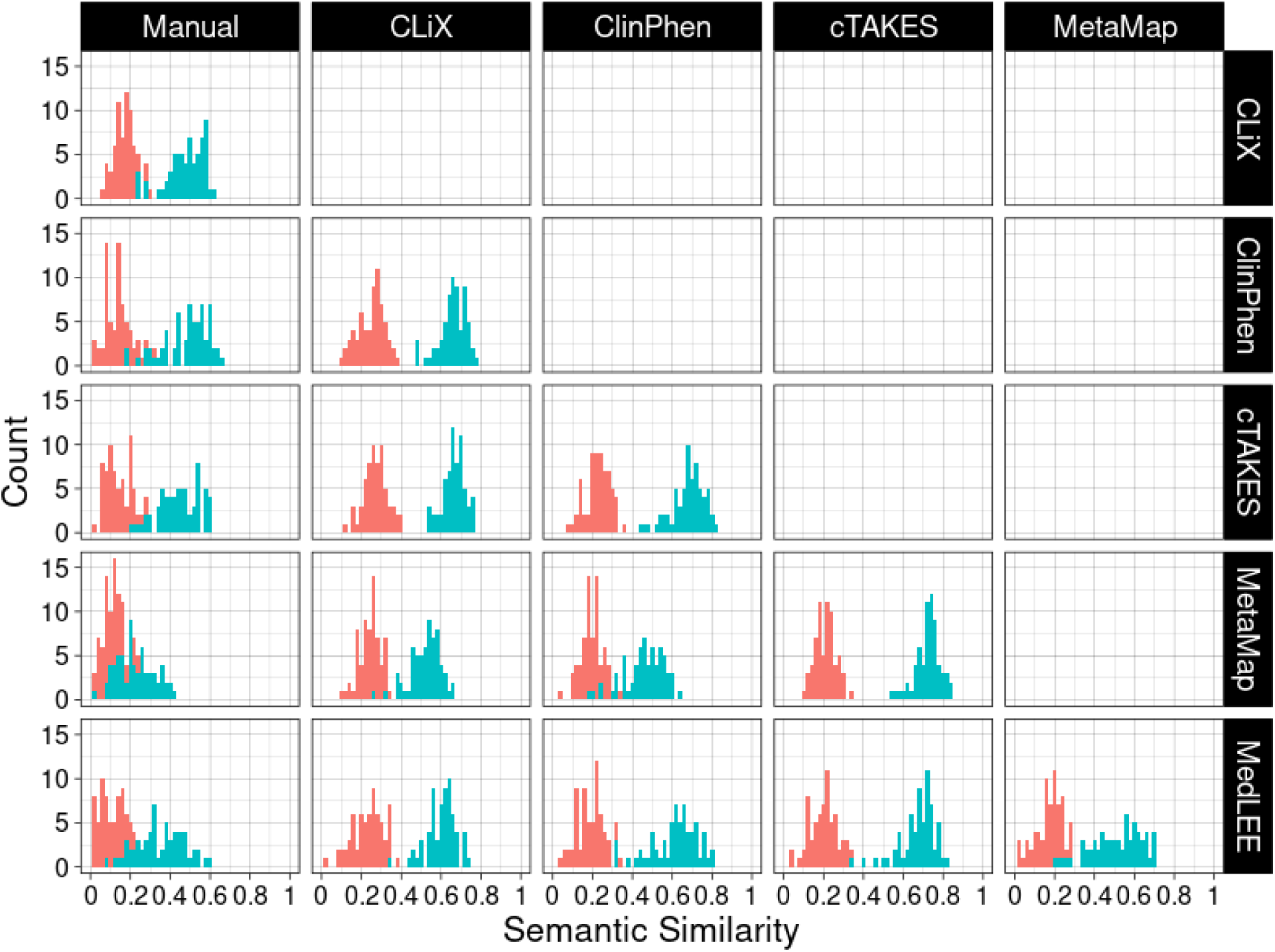
HPO term sets produced by different CNLP tools share robust semantic overlap. Histograms depict the distributions of semantic similarity between phenotype term sets. The histograms in the leftmost column represent comparisons between the CNLP-derived term sets and the manually curated term sets. Blue distributions correspond to the similarity coefficients calculated from the real data, while red distributions represent the similarity coefficients obtained from random sampling of the HPO, with the sample sizes matching those of the respective real term sets. The analysis was conducted on term sets from 65 sequenced Utah NeoSeq patients.

The pronounced separation between semantic similarity distributions of real and simulated data suggests the HPO term sets produced by the tools reflects a common underlying phenotypic reality that is being identified to a greater or lesser degree by all the tools. Consistent with this, the difference in mean similarity between real and simulated datasets is highly statistically significant for every tool by Student’s paired T-test, even after multiple test corrections (data not shown). Interestingly, the Manual sets showed the lowest similarity with the CNLP sets, perhaps due to their smaller number of terms, while overall, cTAKES generally showed the highest mean similarity with other CNLP tools. Among the Manual/NLP comparisons, MetaMapLite had the lowest average similarity coefficient (mean: 0.227, min: 0.02, max: 0.429), while ClinPhen (mean: 0.485, min: 0.174, max: 0.663) and CLiX (mean: 0.479, min: 0.238, max: 0.630) had the highest average similarity coefficients. Among the NLP/NLP comparisons, cTAKES and MetaMapLite had the highest average similarity to one another (mean: 0.722, min: 0.543, max: 0.836), while ClinPhen and MetaMapLite had the lowest average similarity (mean: 0.461, min: 0.171, max: 0.634).

#### NLP Sensitivity and Accuracy

Next, we performed an orthogonal test of CNLP tool sensitivity and accuracy by calculating the overlap between the CNLP-generated phenotype sets and two distinct “ground truth” reference sets: expert-generated phenotypes and OMIM disease-associated phenotypes. The expert reference sets constituted the HPO term lists manually curated by physicians for the 65 sequenced University of Utah NICU patients. The OMIM disease reference sets were restricted to the HPO phenotype terms associated with the OMIM disease diagnosis for the subset of 26 WGS-diagnosed Utah NICU patients. For these analyses, a CNLP term is considered a “true positive” if it or any of its parent terms are found in the ground truth set. This approach is justified by the ontological relationship between parent and child terms in HPO, where a child term inherently implies the presence of its parent term(s). Since HPO is structured in a manner where more specific terms (i.e., child terms) represent refined phenotypic descriptions, they subsume the more general terms (i.e., parent terms). For example, having Thrombocytosis (HP:0001894) necessarily indicates the presence of the parent phenotype Abnormal platelet count (HP:0011873). Consequently, the presence of a specific term in the CNLP set reasonably reflects the occurrence of a broader, related phenotype in the ground truth set.

**Additional file 2: Figure S1** shows sensitivity and accuracy distributions for CNLP terms sets compared with physician manual terms and OMIM disease-associated terms. The relative sensitivity among the CNLP tools roughly correlates with the tools’ average term counts (see **Additional file 1: Table S2**), which isn’t surprising; the more terms a tool generates, the more likely it will capture terms in the reference set. CLiX had the highest average sensitivity among the tools (manual terms sensitivity: 63%; OMIM terms sensitivity: 21%), while MetaMapLite had the lowest (manual terms sensitivity: 15%; OMIM terms sensitivity: 9%). The accuracy measure differs from sensitivity by controlling for the variable sizes of the CNLP term sets. Thus, a CNLP tool with high sensitivity may have a low relative accuracy if it has many more terms than another tool with lower sensitivity. ClinPhen had the highest average accuracy among the tools (manual terms sensitivity: 13%; OMIM terms sensitivity: 11%), while MetaMapLite had the lowest (manual terms sensitivity: 2%; OMIM terms sensitivity: 4%). Despite the modest sensitivity and accuracy of these CNLP tools compared to physician manual terms and OMIM disease-associated terms, MPSE and other phenotype-driven clinical diagnostics tools such as GEM^11^, appear to be very robust against “noisy” phenotype data inputs (see next results for justification).

### MPSE flexibly handles input data from a variety of sources

To further evaluate the practical utility of the MPSE algorithm, we conducted a comparative analysis of different CNLP tools and data types as inputs to MPSE. This analysis addresses several critical considerations for the deployment of MPSE in diverse clinical environments. First, we explored the interoperability of MPSE by assessing whether a model trained with data from one CNLP tool could reliably predict outcomes using data generated by a different tool. Second, we investigated the feasibility of using non-phenotype data with MPSE. The overarching purpose of these analyses is to better understand MPSE’s flexibility, robustness, and broader applicability in real-world clinical settings.

#### MPSE performance using different CNLP tools

To determine whether MPSE, trained with data from one CNLP tool, can reliably predict outcomes using data generated by a different CNLP tool, we began with the original MPSE model trained using CLiX-generated phenotype data from the RCHSD cohort. This model was used to make predictions on external phenotype data from the Utah cohort generated with ClinPhen, CLiX, cTAKES, MedLEE, and MetaMapLite. MPSE’s precision and diagnostic yield among top-scoring probands is plotted in panel A of **Figure 2**. Apart from MetaMapLite, all the CNLP tools’ outputs work well when used as inputs for MPSE, a fact made clear by the high recovery rates of sequenced and diagnosed patients compared to choosing patients randomly for WGS (**Figure 2**). If MPSE was used to automatically select a volume of NICU patients for sequencing identical in size to the Utah NeoSeq study (n=65) from among the 1,838 total patients screened, CLiX and ClinPhen would maintain the NeoSeq study’s physician-mediated diagnostic yield (40%) throughout the top 50% of MPSE scores. This finding accords well with our previous publication, which showed high projected diagnostic yields from MPSE prioritization^12^.

**Figure 2.**
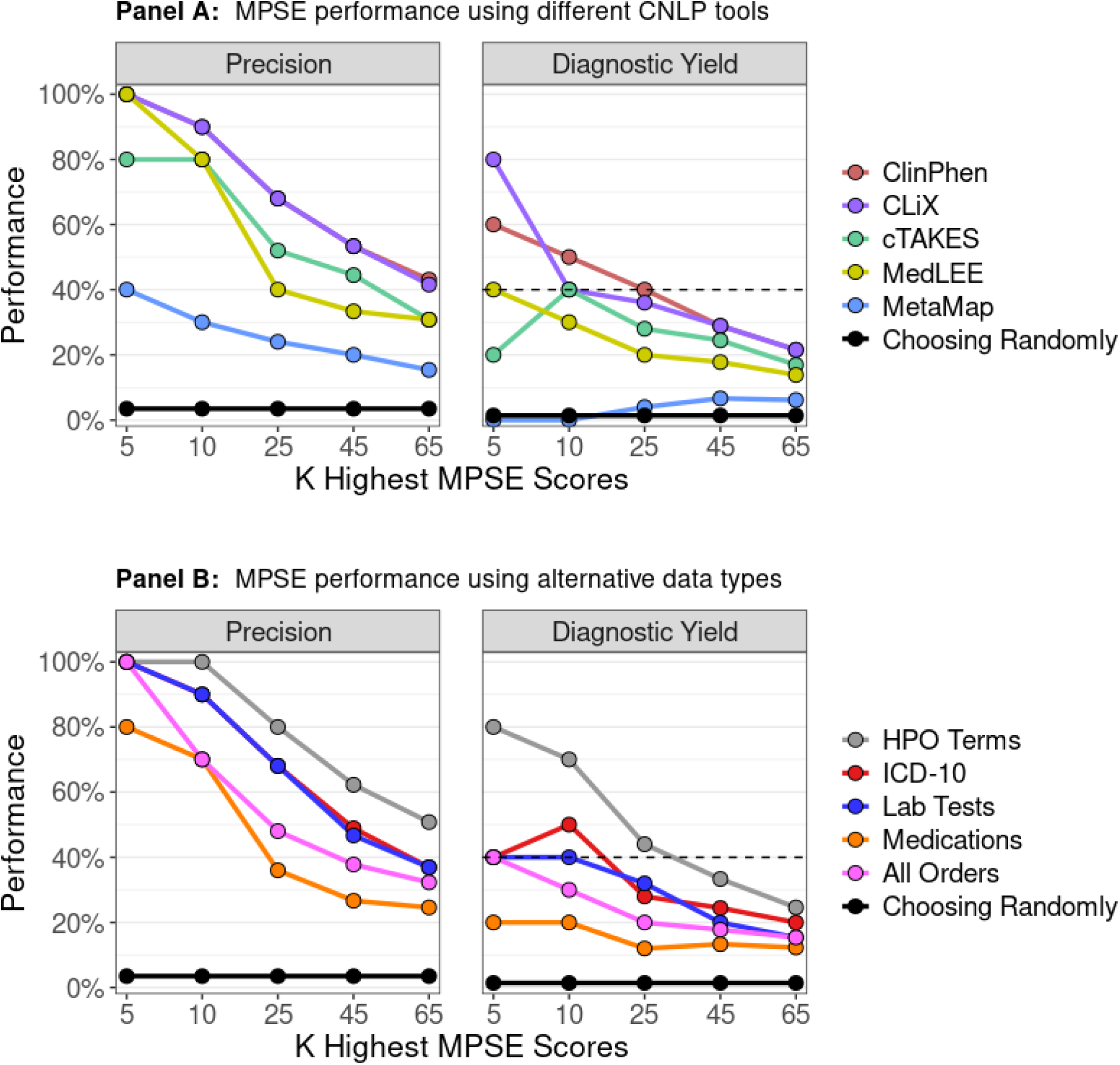
MPSE can ingest different CNLP tool outputs and use alternative data types. Panel A displays MPSE precision rates of patients manually selected for WGS (left) and diagnostic yield for the subset of cases diagnosed by WGS (right) using different CNLP tools. A CLiX-trained MPSE model from the RCHSD cohort was applied to phenotype data from 1,838 University of Utah NICU patients generated by five different CNLP tools. Panel B displays precision and diagnostic yield using MPSE models trained on four alternative data types (diagnosis codes, lab tests, medications, and all orders), compared to the corresponding HPO-based (CLiX) model trained on the same Utah cohort. A solid black reference line in each panel represents the precision or diagnostic yield expected from a model that chooses candidates at random, while the black dashed line in the diagnostic yield graphs (right panels) indicates the NeoSeq study’s 40% total diagnostic yield.

In addition to assessing yield among top-scoring probands, we also calculated cohort-wide performance metrics (see **Additional file 1: Table S3**). After the “native” CLiX data, ClinPhen yields the next-best overall predictions, with an average 20.7-point difference in MPSE score between cases and controls (p=2.2e-14), the highest area under the ROC curve (AUROC=0.91), and the highest area under the PRC curve (AUPRC=0.45). MetaMapLite exhibited the lowest performance, likely caused by the relative dissimilarity between CLiX and MetaMapLite as seen in the low semantic similarity coefficients of CLiX-MetaMapLite term sets in **Figure 1**.

#### MPSE performance using alternative data types

To determine whether non-phenotype data types could be used with MPSE, we tested MPSE models built using diagnosis codes, lab tests, and medications and compared their predictive ability to phenotype-based models. A short description of these data types is given in **Additional file 1: Table S4**, and summary statistics of observation counts for each alternative data type are shown in **Additional file 1: Table S5**. MPSE models trained with alternative data types recovered fewer sequenced cases and diagnostic cases (Figure 2 panel B) among top-scoring probands than a corresponding phenotype-based model but still performed much better than a random model. Among the alternative data types, the ICD-10 based model yielded the best overall predictions, approaching the performance of the CLiX-based model (**Additional file 1: Table S3**). Our analysis suggests that non-phenotype structured data from patient EHRs is less effective than CNLP-derived phenotype data at identifying NICU sequencing candidates, but is a useful and valid substitute for CNLP phenotype descriptions if these are not available. We also compared the utility of combining HPO terms with alternative data types (data not shown) and found that MPSE models trained with combined datasets did not yield better predictions than ones using only HPO terms alone, suggesting that for this application ICD-10 codes, laboratory tests, and medication use do not provide significant orthogonal information to patient phenotypes. However, it should be noted that this analysis was limited to only using the presence/absence of a lab test, medication order, etc. (see Methods) and not the test result or order specifications. An analysis using more precise laboratory and medication data may reveal higher MPSE performance when using these alternative data types.

#### Diagnostic performance using different CNLP tools

Lastly, we assayed the ultimate utility of the CNLP-derived phenotype term sets generated by each tool for clinical molecular diagnostic activities. For these analyses, we used an Artificial Intelligence (AI)-based gene prioritization tool called GEM^11^. Licensed from Fabric Genomics, by both RCHSD and the University of Utah, GEM is a commercial tool that combines HPO-based phenotype descriptions with WES and WGS sequences for rapid, AI-based diagnostic decision support. GEM was used by both RCHSD and the University of Utah for the original diagnosis of every sequenced proband in the datasets analyzed here.

Comparison of GEM’s previously published diagnostic performance to the prospective Utah data reported here provides a unique opportunity both to reexamine GEM’s performance using new, orthogonal data, and to assay the impact of using different CNLP tools on GEM’s diagnostic performance. These data are shown in **Figure 3**. For reference, the original GEM benchmark results using manually curated HPO term sets for 119 RCHSD probands^11^ have been added for ease of comparison. **Figure 3** shows the percentage of diagnosed Utah NeoSeq probands where the clinical molecular diagnostic genotype was reported by GEM among its top 1st, 2nd, 5th, and 10th gene candidates.

**Figure 3.**
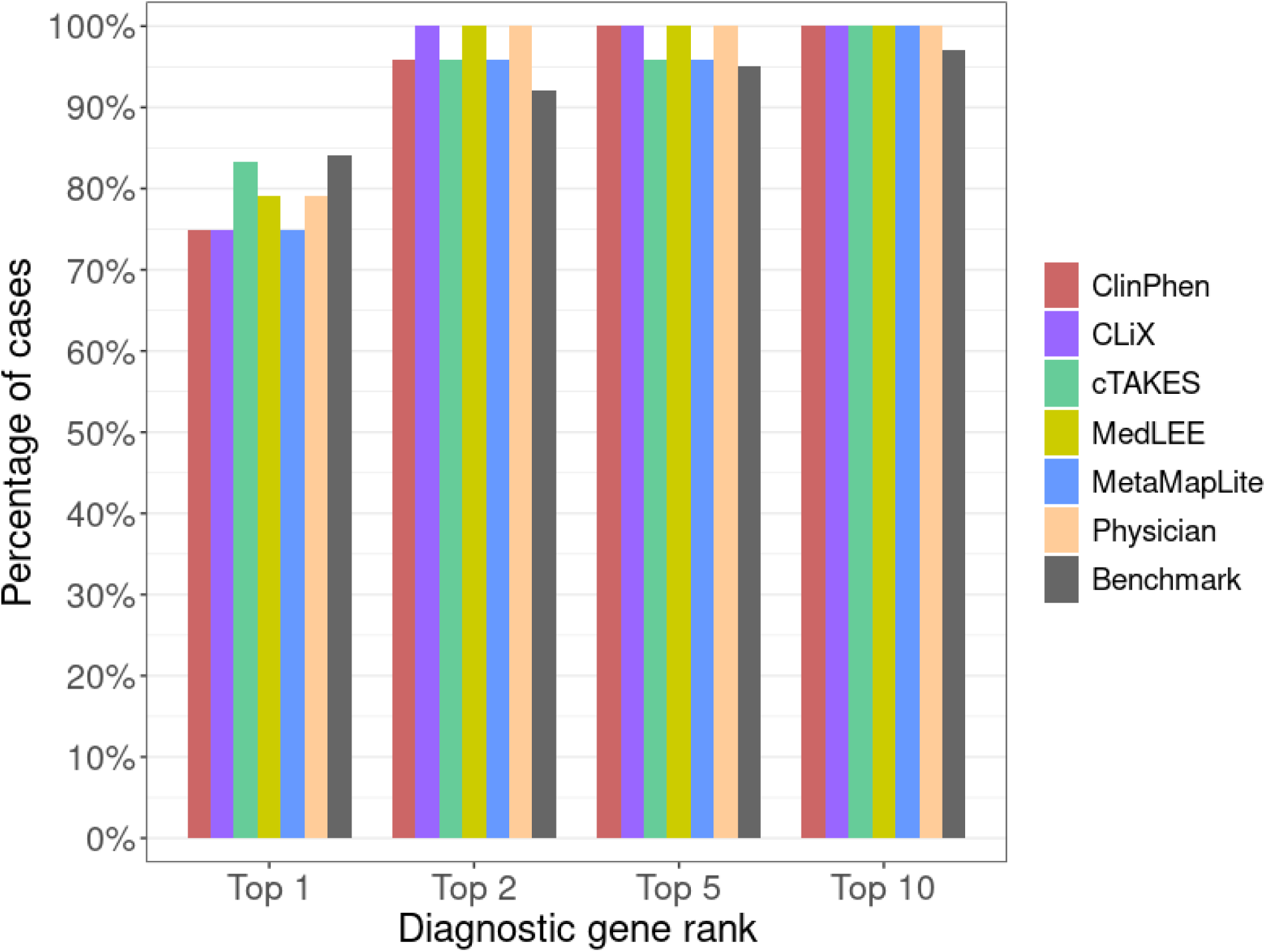
GEM AI performance is agnostic with respect to CNLP tool. Bars show the proportion of diagnosed NeoSeq probands where the true causal genes were identified by GEM among the top 1st, 2nd, 5th, and 10th gene candidates. Each GEM run differed by the input HPO term lists, which were made by extracting phenotypes from patient clinical notes using CNLP (ClinPhen, CLiX, cTAKES, MedLEE, MetaMapLite) or manual physician review. The RCHSD benchmark (n=119 patients) results from the original GEM paper are included for reference (redrawn with authors’ permission).

Two major conclusions emerge from these data. First, diagnostic performance using CNLP-derived HPO data, regardless of the tool used to generate them, is nearly identical to the performance obtained using manual term sets, and second, the results shown here are highly concordant with the original published benchmarking results^11^. Moreover, the University of Utah data provides an entirely prospective orthogonal dataset, demonstrating transportability. These results demonstrate a powerful synergy between the MPSE approach and GEM. Namely, that the same HPO term sets used by MPSE for WGS prioritization can be directly consumed by GEM for downstream diagnoses. Moreover, **Figure 3** makes it clear that GEM can use HPO term sets, manually curated or CNLP derived, regardless of tool, without compromising downstream diagnostic accuracy.

### MPSE can identify patients who would benefit from WGS within the first 48 hours of NICU admission

Our initial work and that presented here has demonstrated MPSE’s ability to accurately identify sequencing candidates by aggregating information from the entirety of the patient’s NICU stay. These findings underscore the tool’s effectiveness in a retrospective context, using all notes up to the date at which the attending physicians place the order for WGS. However, the ultimate test of value lies in validating the real-time utility of MPSE. Early identification of patients who would benefit from WGS, as soon as possible after NICU admission, could significantly enhance care, enabling earlier disease diagnosis and more timely interventions^2,14,15^.

To measure MPSE’s real-time utility, we calculated daily MPSE scores for each patient in our Utah cohort using only HPO terms extracted from clinical notes present in the EHR at 24-hour intervals, beginning at the moment of their admission. Thus, each patient had a series of MPSE scores for each day spent in the NICU from admission to discharge. Longitudinal MPSE scores for patients who received a molecular diagnosis by WGS (diagnostic), those for whom WGS did not identify a molecular diagnosis (non-diagnostic), and patients who were not selected for WGS (unsequenced) are summarized in **Additional file 1: Table S6** and plotted in **Figure 4** to help visualize the change in MPSE score over time among these groups. By the end of the first day (0 to 24 hours) in the NICU, both diagnostic and non-diagnostic sequenced cases had statistically significantly higher MPSE scores than did those who were not selected for sequencing (unsequenced mean: -48.4; diagnostic mean: -32.1, p=1.4e-5; non-diagnostic mean: -28.2, p=9.3e-6). Additionally, diagnostic cases have significantly higher average MPSE scores than non-diagnostic sequenced cases beginning 48 hours post-admission (non-diagnostic mean: -24.7; diagnostic mean: -9.0; p=0.018) and continuing thereafter.

**Figure 4.**
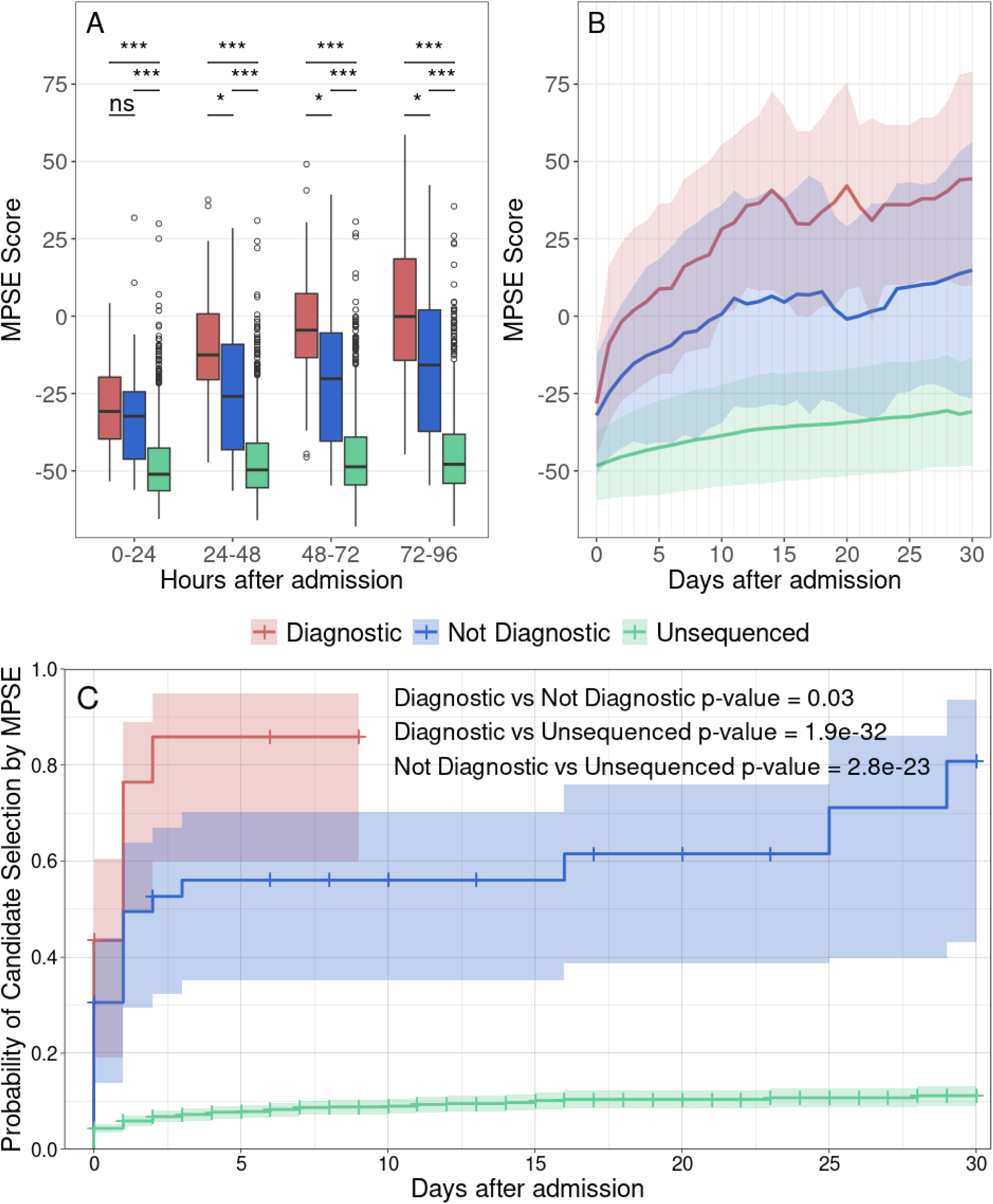
MPSE enables automated WGS candidate identification within the first 24 hours in the NICU. Panel A shows MPSE score distributions across the first 96 hours in the NICU for diagnostic (red) and sequenced but non-diagnostic (blue) patients as well as unsequenced NICU patients (green). Diagnostic and non-diagnostic sequenced patients had significantly higher MPSE scores than unsequenced patients beginning 0 to 24 hours after admission. Diagnostic patients had significantly higher MPSE scores than non-diagnostic patients beginning at 48 hours post-admission. Boxplot comparison significance levels: *** (p < 1e-5); * (p < 0.05). Panel B shows MPSE score trajectories for these groups across the first 30 days in the NICU. Solid lines show the mean MPSE score per group and the shaded regions cover one standard deviation from each mean. Panel C shows the probability of at-risk patients being classified as a WGS candidate by MPSE (i.e., MPSE score > 2 standard deviations above the unsequenced mean score) as a function of time. Cox proportional hazards regression analysis confirmed the significantly increased rate of MPSE candidate selection for diagnostic and non-diagnostic patients selected for WGS compared to unsequenced patients, with hazards ratios of 18.8 (95% CI 11.6-30.6; p=1.9e-32) and 9.8 (95% CI 6.2-15.3; p=2.8e-23) respectively. Diagnostic patients were also selected by MPSE at a higher rate than sequenced but non-diagnostic patients (HR=2.0; 95% CI 1.1-3.9; p=0.03).

In addition to initial differences in MPSE scores between unsequenced, non-diagnostic, and diagnostic patients, there are also significant differences in the daily change in MPSE score (day-N delta) between these groups. Both diagnostic and non-diagnostic sequenced cases saw greater average day-N delta than unsequenced controls throughout the first 30 days post-admission (**Additional file 1: Table S6**). Importantly, the greatest difference in MPSE score increase was observed for the day-one delta, i.e. the change in MPSE score during the first 24 hours post-admission, with average diagnostic MPSE score rising by 18.4 points, average non-diagnostic MPSE score rising by 7.7 points, and average unsequenced MPSE score rising by only 2.7 points. These day-one delta differences were statistically significant for each subgroup comparison (diagnostic vs unsequenced p-value = 0.0015; non-diagnostic vs unsequenced p-value = 0.004; diagnostic vs non-diagnostic p-value = 0.027). The observed MPSE score trajectories clearly show a marked separation between unsequenced controls, non-diagnostic cases, and diagnostic cases that appears immediately after admission and only grows more pronounced over time.

To gain further insight into the temporal dynamics of MPSE’s predictive capabilities across the NICU stay, we estimated the proportional risk of being identified as a WGS candidate by MPSE using Cox proportional hazards regression analysis^37^. The estimated probability and 95% CI of candidate selection by MPSE for diagnostic, non-diagnostic, and unsequenced patients is plotted in Panel C of **Figure 4**. Using a rule-of-thumb MPSE score threshold (calculated individually for each day) of 2 standard deviations above the mean score of unsequenced control patients, diagnostic patients were flagged by MPSE at a significantly higher frequency and speed than both unsequenced patients (HR=18.8, p=1.9e-32) and non-diagnostic sequenced patients (HR=2.0, p=0.03). At 72 hours post-admission, MPSE had already flagged 83% (19 of 23) of diagnostic patients, 50% (18 of 36) of non-diagnostic sequenced patients, and only 6% (113 of 1773) of unsequenced patients. Furthermore, by 9 days post-admission all diagnostic patients had been flagged by MPSE or were censored as a result of death or NICU discharge, highlighting the speed at which MPSE was able to determine correct clinical action for this group of patients. **Additional file 1: Table S7** contains the daily score threshold, the number of candidates assessed by MPSE, and the cumulative number of patients who reached the score threshold as part of this longitudinal analysis.

Taken together, these results in **Figure 4**, **Additional file 1: Table S6, and Additional file 1: Table S7** show that MPSE can be used not only to identify patients who would likely benefit from WGS, but also which of those patients will ultimately prove diagnostic for Mendelian disease. Moreover, MPSE can do so within the first 24 to 48 hours post NICU admission. These findings make clear MPSE’s value as a real-time tool and how MPSE could be used to improve cost savings^2,3,14,15^ and the timeliness and effectiveness of care. The consistent performance of MPSE over the first 30 days post-admission argues for its utility as a monitoring tool throughout the entirety of a patient’s NICU stay.

## Conclusions

We previously demonstrated that an MPSE-based automated pipeline for prioritizing acutely ill infants for whole genome sequencing can meet or exceed diagnostic yields obtained by time-consuming manual review of clinical notes and histories. Our work here serves to expand on those original findings. Here we have shown that MPSE’s performance is largely agnostic with respect to upstream CNLP tools. Moreover, we show that structured EHR data, such as ICD diagnosis codes, can provide an effective alternative for prioritizing patients for WGS in health settings where access to clinical notes and NLP pipelines is problematic. These two features of MPSE combine to greatly lower the IT burden for deployment.

Our longitudinal analyses demonstrate that MPSE can identify those children most likely to benefit from WGS within the first 48 hours of admission to the NICU, a critical window for maximally impactful care. Moreover, the consistent performance of MPSE over the first 30 days post-admission argues for its utility as a monitoring tool throughout the entirety of a patient’s NICU stay.

Finally, we have also shown that the same HPO term sets used by MPSE for prioritization for WGS, regardless of the CNLP tool generating them, can be directly consumed by the AI tool GEM for downstream molecular diagnoses, further speeding and facilitating personalized care. These results collectively demonstrate that MPSE provides fast, flexible, and highly scalable means for prioritizing critically ill newborns for whole genome sequencing.

The American College of Medical Genetics and Genomics (ACMG) 2021 recommends clinical genome sequencing as a first or second-tier test for infants with one or more congenital anomalies^25^. We compared the efficiency of using this ACMG guideline to using MPSE for patient selection. The MPSE diagnostic yield at day 10 post-admission was 11% (19 diagnostic patients out of 171 selected). The ACMG guideline-based diagnostic yield was 2% (21 diagnostic patients out of 1029 selected). These results suggest MPSE can provide a 5.5-fold enrichment in diagnostic rate compared to ACMG criteria alone while achieving essentially the same number (19 vs. 21) of diagnosed children.

Despite overwhelming clinical evidence that NICU and PICU genome sequencing saves lives and reduces costs^3,14,15,25^, several barriers still hinder its broad adoption. One significant obstacle is reimbursement for testing by payers^38–40^. Broader, more inclusive eligibility criteria simplify the candidate selection process, but increase cost-burden and decrease diagnostic yield. This can result in payer hesitancy to reimburse WGS, especially for negative results. While more stringent selection criteria can decrease cost-burden and increase diagnostic rates, they also increase the time-burden of candidate assessment. Collectively, our results demonstrate how MPSE can provide means to overcome the limitations of using rule-based eligibility criteria, democratizing WGS in the NICU and PICU. Moving forward, we will evaluate the benefits of large-scale MPSE implementation across the Intermountain West (funded by a grant from the Warren Alpert Foundation) and explore secondary applications of MPSE in assessing eligibility for reimbursement.

## Data Availability

All data produced in the present study are available upon reasonable request to the authors. MPSE source code, pre-trained models, documentation, and synthetic datasets are available to the public on GitHub (https://github.com/Yandell-Lab/MPSE).

