## Additional file 2 for "MPSE provides fast, flexible, and efficient means to identify newborns who will benefit from whole genome sequencing within the first 48 hours of NICU admission"

**Supplemental Figure S1**. CNLP tool sensitivity and accuracy.

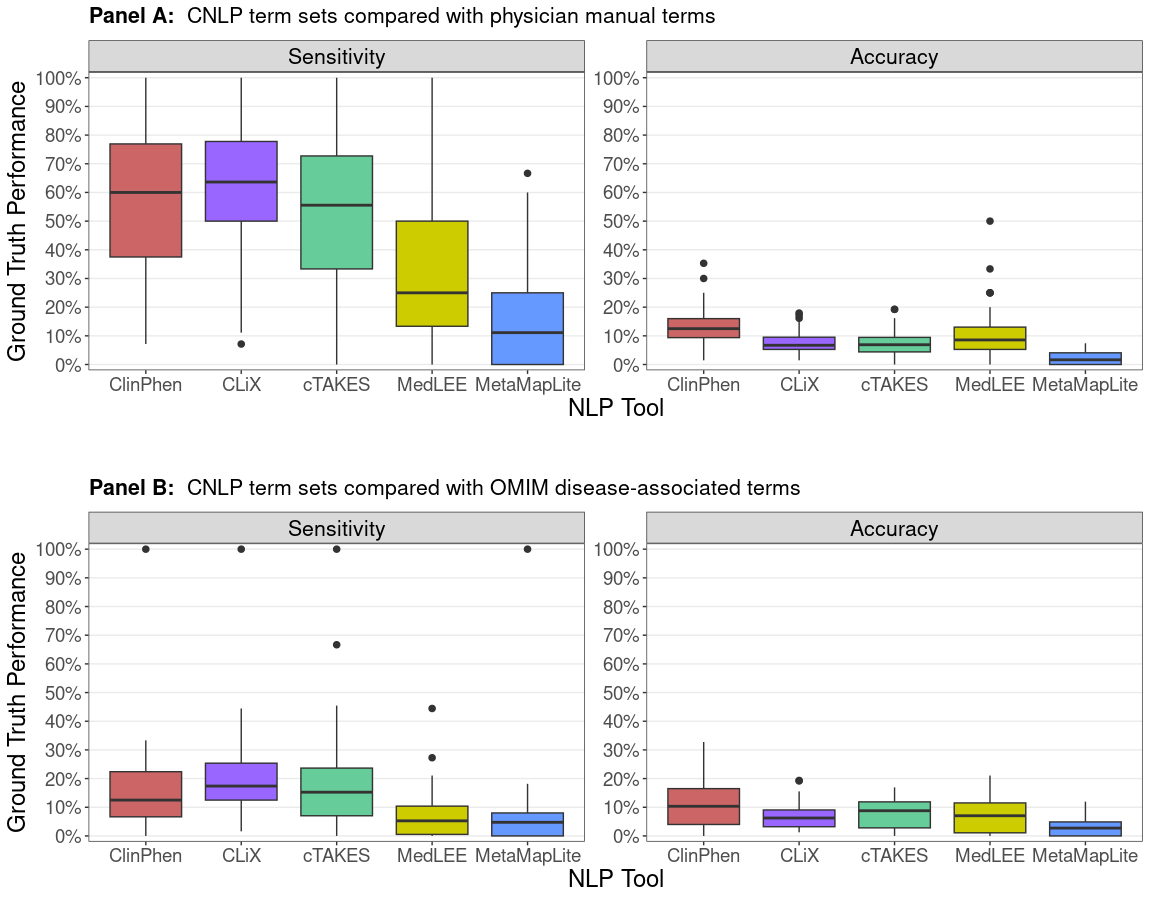

**Figure S1.** **CNLP tool sensitivity and accuracy.** Box plots show the sensitivity and accuracy of CNLP term sets when compared with physician-generated “ground truth” terms (Panel A) and OMIM disease associated terms (Panel B). Data comes from 65 sequenced Utah NeoSeq patients. OMIM term lists for the 26 WGS-diagnosed NeoSeq patients were gathered from the HPO phenotype annotations of OMIM diseases. Sensitivity is the ratio of true positives to the number of “ground truth” terms. Accuracy is the ratio of true positives to the number of unique terms in the CNLP set.
